# CSF proteogenomics implicates novel proteins and humoral immunity in Alzheimer’s disease risk

**DOI:** 10.1101/2025.09.21.25336297

**Authors:** Aliza P. Wingo, Yue Liu, Zhen Mei, Michael Wang, Selina M. Vattathil, Ekaterina S. Gerasimov, Anantharaman Shantaraman, Fang Wu, Duc M. Duong, Edward J. Fox, Alzheimer’s Disease Neuroimaging Initiative, David A. Bennett, Allan I. Levey, Nicholas T. Seyfried, Thomas S. Wingo

**Affiliations:** VA Northern California Healthcare System, Sacramento, CA, USA; Department of Psychiatry, University of California, Davis, Sacramento, CA, USA; Department of Neurology, University of California, Davis, Sacramento, CA, USA; Department of Biochemistry, Emory University School of Medicine, Atlanta, GA, USA; Center for Neurodegenerative Disease, Emory University School of Medicine, Atlanta, GA, USA; Rush Alzheimer’s Disease Center, Rush University Medical Center, Chicago, Illinois, USA; Goizueta Alzheimer’s Disease Research Center, Emory University School of Medicine, Atlanta, GA, USA; Department of Neurology, Emory University School of Medicine, Atlanta, GA, USA; Alzheimer’s Disease Research Center, University of California, Davis, Sacramento, USA

## Abstract

Identifying causal genes at Alzheimer’s Disease (AD) GWAS loci requires tissue-relevant functional genomic data. Cerebrospinal fluid (CSF) interfaces directly with the brain and can be obtained from living individuals, providing a potential proxy for brain protein biology at scale; however, large-scale CSF proteogenomic studies have been limited to affinity-based platforms. Here, we used mass spectrometry to profile CSF proteins in 1,005 participants measuring 2,961 proteins, including 1,066 proteins not captured by other platforms. We mapped protein quantitative trait loci (pQTLs) in CSF, compared them with brain and plasma pQTLs, and integrated them with AD GWAS results. We identified 1,417 *cis* and 130 *trans* pQTLs, and uncover 24 candidate causal proteins – 10 novel, including 5 detected only by mass spectrometry. These proteins implicate lysosomal function, neurovascular remodeling, and immune response. Notably, three immunoglobulins (IGHG2, IGHV2-70, IGHV3-72) link humoral immunity to AD risk for the first time.

## Introduction

Genome-wide association studies (GWAS) have identified many disease-associated genomic regions, yet for most loci, the underlying biological mechanisms remain unknown. A prevalent hypothesis supported by considerable evidence^1^ is that GWAS variants influence disease by altering gene expression. Functional genomics tests this hypothesis by mapping quantitative trait loci (QTLs) – genetic variants that regulate molecular phenotypes like gene expression (eQTLs) or protein abundance (pQTLs) – and integrating them with GWAS results. Integration of QTL data with GWAS can reveal whether disease associations at a locus are explained by effects on gene expression. Approaches like Summary-based Mendelian Randomization (SMR)^2^ and colocalization^3,4^ can identify genes whose genetically regulated expression is consistent with a causal role in disease from those with coincidental overlap due to linkage disequilibrium (LD). These integration methods point us to future testable hypotheses about molecular function and therapeutic targets for disease.

Tissue-specific functional genomic studies provide important insight into GWAS results because genetic regulation of gene expression varies across tissues, cell types, and developmental stages. While plasma proteomic studies have profiled tens of thousands of individual samples and provide exceptional power to resolve pQTLs^5,6^, plasma likely offers limited insights into brain-specific processes. Conversely, functional proteomics of brain tissue have illuminated novel molecular mechanisms in Alzheimer’s disease and other neurological conditions^7–10^, but the sample sizes are constrained by availability^9,11^. Cerebrospinal fluid (CSF) represents an intermediate for investigating brain-related traits and diseases because it directly interfaces with brain tissue, can reflect neurodegenerative disease processes, and can be obtained from living individuals.

Large-scale functional genomic studies of CSF proteomics to date have relied predominantly on affinity-based platforms – either aptamer-based (SomaLogic)^9,12,13^ or antibody-based (Olink)^14^ – that target predefined proteins. By contrast, bottom-up mass spectrometry-based (MS) proteomics enables untargeted protein identification and quantification, revealing proteins not captured by existing affinity panels.

Here, we present a large-scale functional genomics study of CSF using tandem mass tag mass spectrometry (MS) based proteomics in 1,005 individuals (**Figure 1**). Using an untargeted approach, we identified 2,961 proteins, including 1066 not measured in prior affinity-based CSF studies. We performed *cis* and *trans* pQTL mapping, sex-stratified pQTL testing, and fine mapping of *cis* pQTLs that allow us to compare MS-based CSF pQTLs with MS-based brain pQTLs and affinity-based CSF and plasma pQTLs. Given the proximity of CSF to brain and ability to collect CSF in living participants, we compared proteins differentially expressed between control and Alzheimer’s disease (AD) dementia in CSF and brain. Finally, integrating CSF pQTLs with AD GWAS using proteome-wide association study (PWAS)^4^ and SMR Mendelian randomization^2^ revealed 24 putatively AD causal proteins, including 10 new and 5 of these were detected only by mass spectrometry. Together, these findings provide novel insights into AD pathogenesis and link humoral immunity in AD risk for the first time.

**Figure 1.**
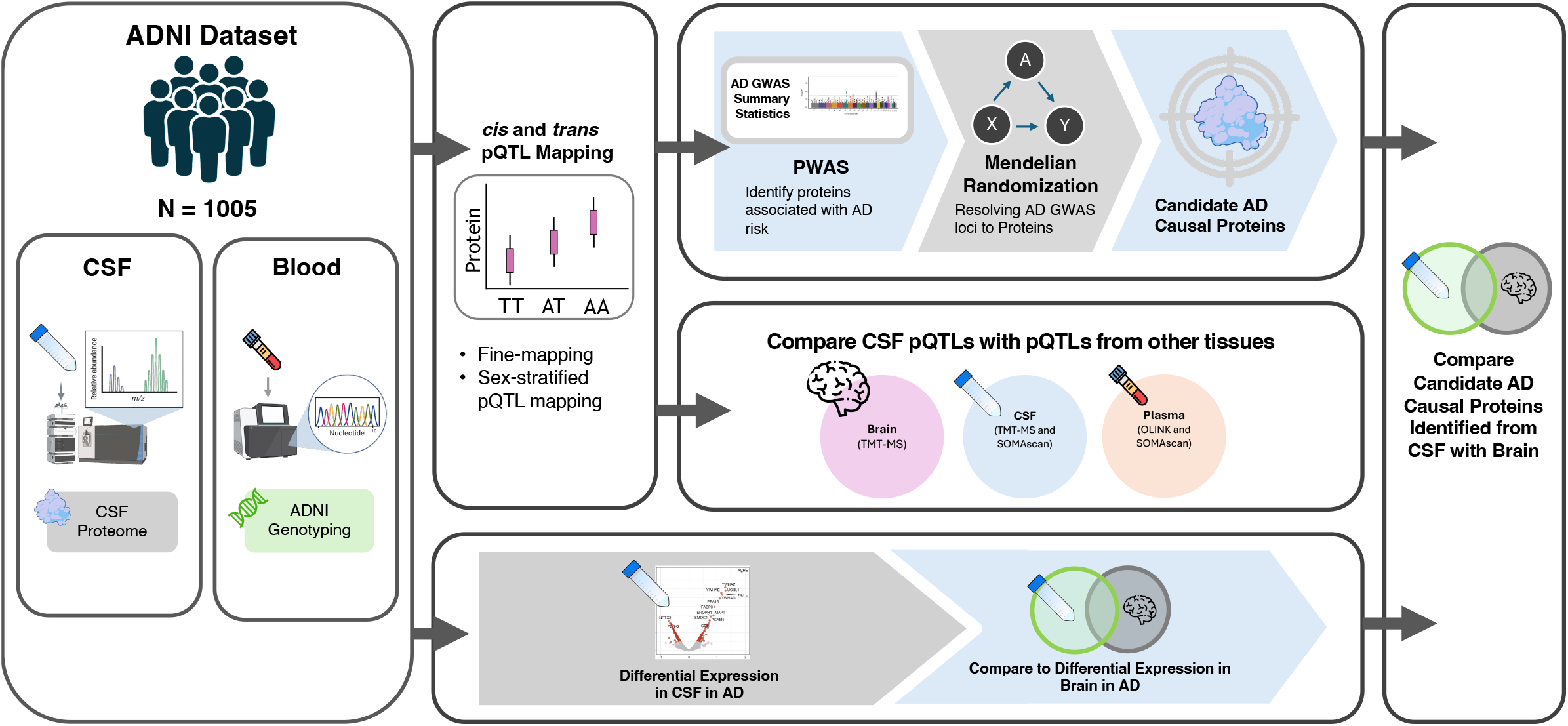
Study Overview. Study participants were individuals from ADNI who donated cerebrospinal fluid (CSF) and had available genotyping. Mass spectrometry (MS) was used to generate CSF proteomes and those were used to perform *cis* and *trans* protein quantitative trait locus (pQTL) mapping. Fine mapping was performed on the *cis* pQTLs. Sex-specific pQTLs were also identified. MS-based proteomic pQTLs were compared with published studies using CSF, brain, and plasma tissue and MS and affinity-based approaches for measuring the proteome. The CSF proteome was compared with brain proteome in differential analysis of AD dementia. Finally, the CSF pQTLs were used to infer AD causal proteins that were compared with results of brain illustrating CSF-specific findings.

## Results

### A. CSF *cis* pQTL mapping and fine mapping

We profiled CSF proteomes of 1,005 ADNI participants of European ancestry, among whom 45% were female and the mean age was 73.1 years (Table S1). A total of 2,961 CSF proteins were considered for *cis* pQTL mapping after quality control and regressing out effects of proteomic sequencing batch, age, clinical diagnosis, and surrogate variables. We defined *cis* pQTLs as SNPs located within 500 Kb up or downstream of the corresponding protein-coding genes that have SNP-protein pair association at *p* < 5 × 10^−8^. We identified 67,296 *cis* pQTLs, and, after clumping for linkage disequilibrium (LD) r^2^<0.1 to keep approximately independent sites, there were 1,417 index pQTLs for 654 unique genes, with a median of 1 SNP per gene (range 1-13, interquartile range [IQR] of 2; Table S2-3). The inflation factor, *λ*, for the pQTL mapping was 1.002 (range of 0.975 – 1.028), indicating no significant inflation. Fine mapping with SuSiE^15^ identified 1,092 credible sets for 739 proteins. There was a median of 6 SNPs per credible set and imposing posterior inclusion probability >50% yielded 457 putative causal pQTLs for 346 unique proteins (Table S4).

To determine whether sex has an influence on the genetic regulation of protein expression in CSF, we performed sex-stratified *cis* pQTL mapping. We detected 30,383 *cis* pQTLs in males and 38,058 *cis* pQTLs in females. Among the cis pQTLs from the joint and sex-stratified pQTL mapping, we examined whether they interact with sex. We identified 2 pQTLs with significant interaction with sex (i.e., *sex* × *pQTL* term had FDR <0.05; Table S5). These sex-diverged CSF pQTLs were associated with expression of pregnancy zone protein (PZP).

### B. CSF *trans* pQTL mapping

CSF *trans* pQTLs were defined as SNPs located outside of the ± 500 kb window of a gene and associated with its protein abundance at *p* < 1.70 × 10^−11^ (*p* < 5 × 10^−8^ ÷ 2931 *genes*). With this threshold, there were 8,411 *trans* pQTLs, corresponding to 130 index *trans* pQTLs (after clumping at LD r^2^ <0.1) for 94 unique genes (*trans* pGene; **Figure 2**; Table S6-8). Four proteins (CPN2, CTSS, LAMA2, and LAMB1) had *trans* pQTLs located on two different chromosomes. The *APOE* locus was the *trans* pQTL for SMOC1 and the 14-3-3 family of proteins (YWHAE, YWHAG, and YWHAZ), which are proteins implicated in early AD pathophysiology and neurodegeneration^16^ (Table S8). However, stratification by AD status or amyloid/tau biomarkers eliminated these associations, suggesting the trans effect is confounded by disease state rather than reflecting direct genetic regulation (Table S9). A recent aptamer-based CSF pQTL study^13^ also identified APOE as a *trans* pQTL for these proteins but reported a substantially larger number of APOE-associated proteins (335 vs. 4), likely reflecting differences in covariate adjustment. Among the 55 *trans* pGenes identified in both this and the CSF aptamer-based study^13^, 33% (18 of 55) shared a *trans* pQTL in linkage disequilibrium (LD) of *r*^2^ > 0.3, suggesting modest cross-platform agreement.

**Figure 2.**
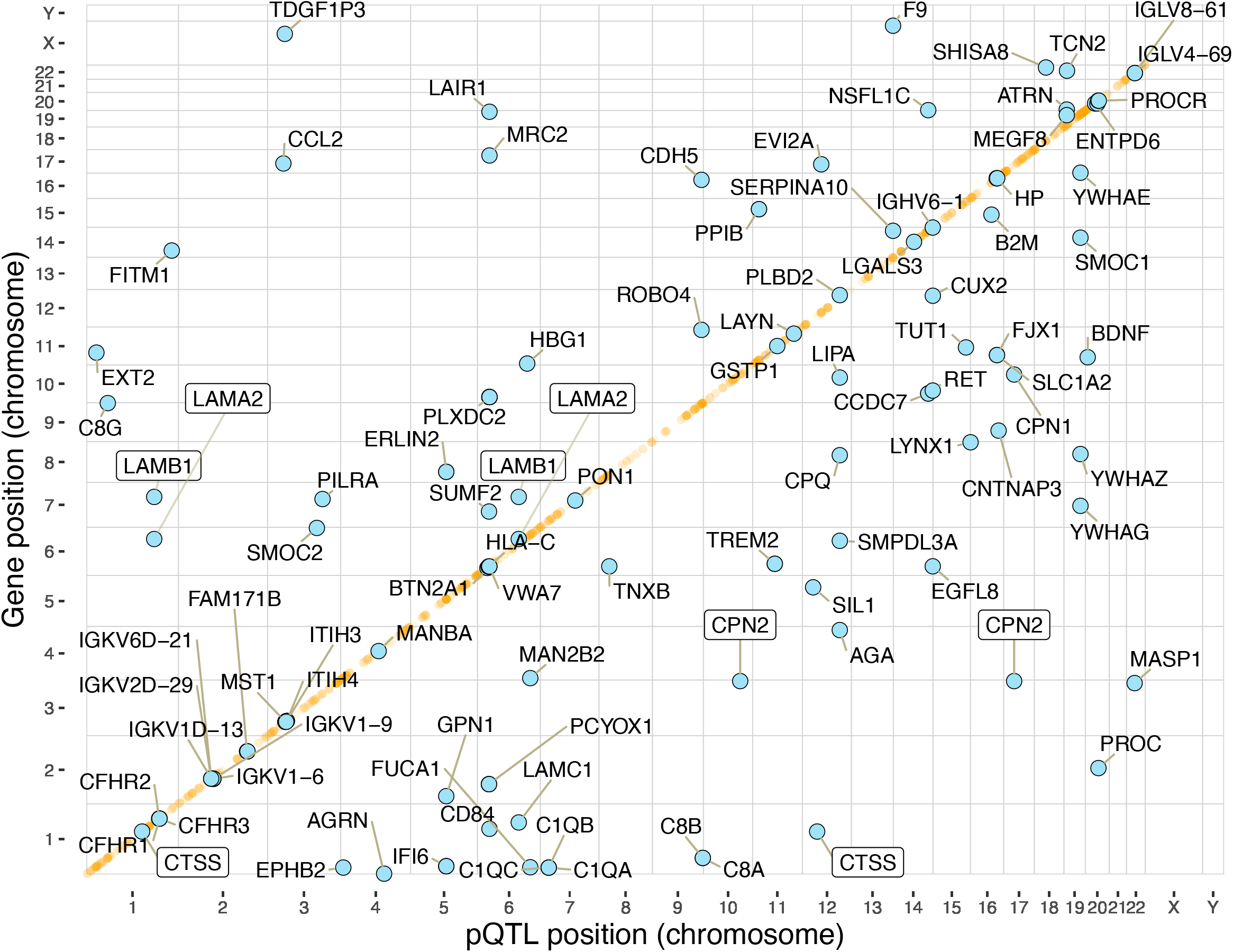
Two-dimensional Manhattan plot for MS-based CSF pQTLs. There are 1,417 index cis pQTLs and 130 index trans pQTLs plotted. The x-axis is the genomic position of the pQTL SNP, and the y-axis is the gene position for the tested protein. The *cis* pQTLs are shown in orange while *trans* pQTLs are shown in blue with their corresponding protein labels. Boxes around gene labels indicate proteins with more than one pQTL on separate chromosomes.

### C. Cross tissue and platform pQTL comparisons

We compared CSF pQTLs with published pQTLs from brain, CSF, and plasma profiled using different technologies: TMT-MS (brain)^10^, SOMAscan (CSF^13^ and plasma^5^), and Olink (plasma^6^) (**Figure 3a**). Between MS- and aptamer-based CSF proteomes, 1,895 proteins overlapped and 1,066 were measured only by MS (**Figure 3b**). Among these MS-only proteins, 181 had cis pQTLs and 32 had trans pQTLs. Between CSF and brain MS-based proteomes, 2,200 proteins were shared, representing 74% of the CSF proteome (**Figure 3c**; Table S10). Between CSF and plasma, the MS-based CSF proteome measured 848 proteins not profiled in either CSF or plasma by other technologies (**Figure 3d**), again highlighting the additional insight into biology that MS-based CSF proteome can provide.

**Figure 3.**
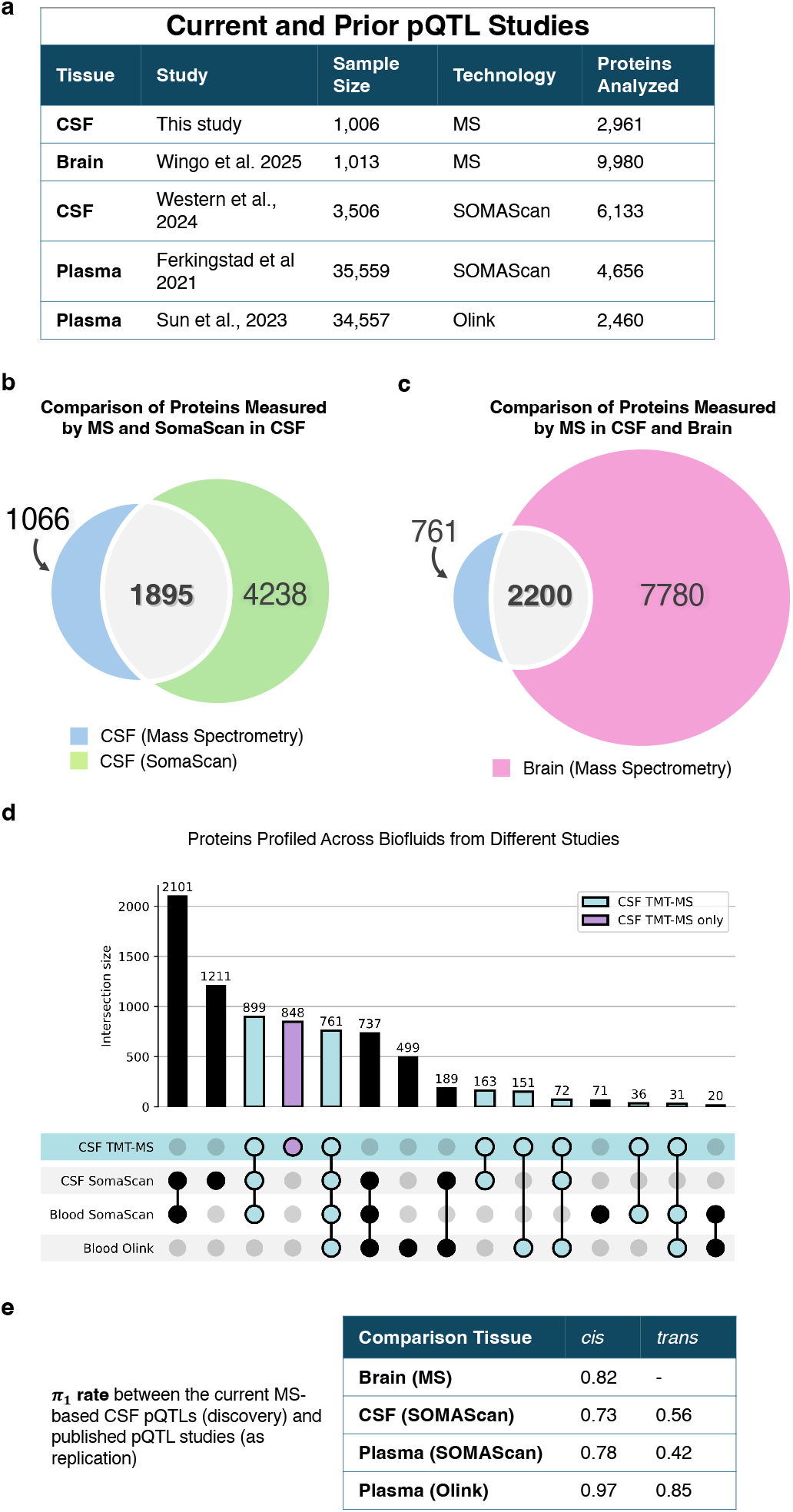
Comparison of MS-based CSF proteomic profiles and pQTLs across different tissues and technologies. **a**. Overview of current and prior pQTL studies included in the comparative analysis, showing tissue type, sample size, proteomic technology, and number of genes analyzed. **b**. Comparison of CSF gene products measured by MS versus aptamer-based technology (in Western et al 2024) shows 1,895 proteins measured in common. **c**. Comparison of gene products measured by MS in CSF (current study) versus brain tissue (Wingo et al. 2025) revealed 2,200 shared genes. **(d)** Comparison of proteins profiled across CSF and plasma in MS- and affinity-based proteomic studies. Bars represent the number of genes detected in each unique combination of studies, with filled circles indicating which studies contribute to each intersection. The blue color indicates proteins measured by CSF TMT-MS and other platforms and purple indicates proteins only profiled by CSF TMT-MS alone. **e**. Replication rates (π_1_) between the current MS-based CSF pQTLs and published pQTL studies for proteins profiled between the two technologies. Results are stratified by *cis* and *trans* pQTLs.

To assess replication, we estimated *π*_1_ rates^17^ between our CSF pQTLs and published pQTL studies across tissues and platforms. Cross-platform *π*_1_ rates were moderate to high for cis pQTLs (0.73–0.97) but lower for trans pQTLs (0.42–0.85; **Figure 3e**). Colocalization analysis found that 35% of CSF pGenes (159 of 445) shared cis pQTLs with brain (Table S11).

### D. Identifying AD candidate causal proteins in CSF

To identify proteins in CSF that are consistent with a causal role in AD, we integrated the largest AD GWAS results^18^ with CSF proteomic and genetic data via a proteome-wide association study (PWAS) using FUSION^4^. In the PWAS, there were 819 proteins with heritable abundance, of which 41 showed genetic evidence for association with AD (Table S12). After Mendelian randomization using SMR/HEIDI^2^, 24 proteins remained significant, suggesting a causal role in AD (**Table 1**; Table S13).

**Table 1.**
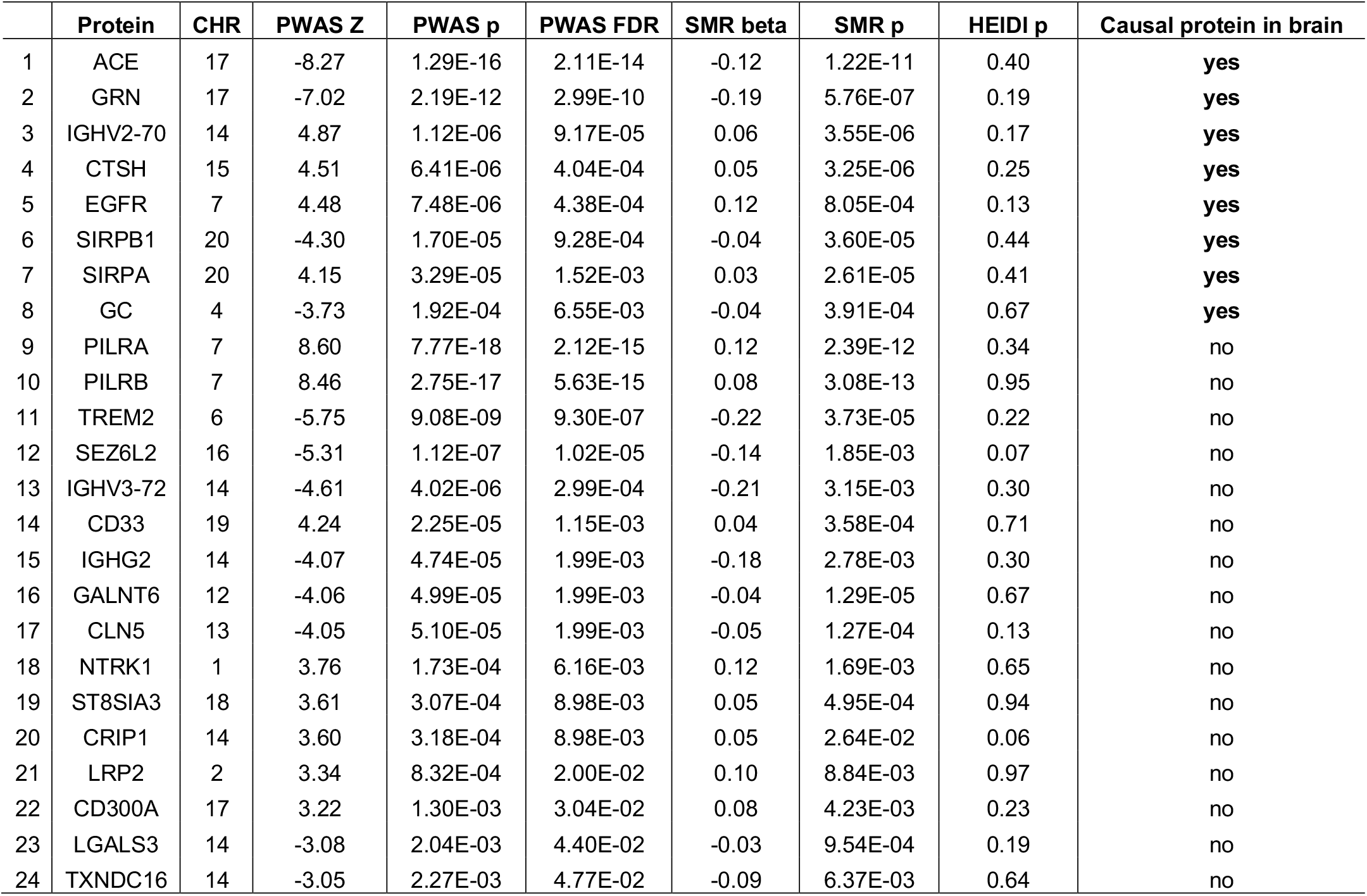
Candidate AD causal proteins identified in CSF (n = 24) Listed are proteins found to be significant in the PWAS of AD and SMR/HEIDI of AD using proteogenomic data from CSF (n = 24). Significance was defined as meeting all the following criteria: PWAS FDR p<0.05, SMR p<0.05; HEIDI p >0.05; SMR beta and PWAS Z have consistent direction of association. Candidate causal proteins in brain were obtained from integrating the same AD GWAS with brain proteogenomic data via PWAS of AD and SMR/HEIDI of AD based on similar criteria. These analyses were previously performed and published by Wingo et al, 2025^7^. Rows were sorted by whether the candidate causal proteins identified in CSF are also candidate causal proteins identified in brain and by PWAS p-values.

|  | Protein | CHR | PWAS Z | PWAS p | PWAS FDR | SMR beta | SMR p | HEIDI p | Causal protein in brain |
| --- | --- | --- | --- | --- | --- | --- | --- | --- | --- |
| 1 | ACE | 17 | -8.27 | 1.29E-16 | 2.11E-14 | -0.12 | 1.22E-11 | 0.40 | yes |
| 2 | GRN | 17 | -7.02 | 2.19E-12 | 2.99E-10 | -0.19 | 5.76E-07 | 0.19 | yes |
| 3 | IGHV2-70 | 14 | 4.87 | 1.12E-06 | 9.17E-05 | 0.06 | 3.55E-06 | 0.17 | yes |
| 4 | CTSH | 15 | 4.51 | 6.41E-06 | 4.04E-04 | 0.05 | 3.25E-06 | 0.25 | yes |
| 5 | EGFR | 7 | 4.48 | 7.48E-06 | 4.38E-04 | 0.12 | 8.05E-04 | 0.13 | yes |
| 6 | SIRPB1 | 20 | -4.30 | 1.70E-05 | 9.28E-04 | -0.04 | 3.60E-05 | 0.44 | yes |
| 7 | SIRPA | 20 | 4.15 | 3.29E-05 | 1.52E-03 | 0.03 | 2.61E-05 | 0.41 | yes |
| 8 | GC | 4 | -3.73 | 1.92E-04 | 6.55E-03 | -0.04 | 3.91E-04 | 0.67 | yes |
| 9 | PILRA | 7 | 8.60 | 7.77E-18 | 2.12E-15 | 0.12 | 2.39E-12 | 0.34 | no |
| 10 | PILRB | 7 | 8.46 | 2.75E-17 | 5.63E-15 | 0.08 | 3.08E-13 | 0.95 | no |
| 11 | TREM2 | 6 | -5.75 | 9.08E-09 | 9.30E-07 | -0.22 | 3.73E-05 | 0.22 | no |
| 12 | SEZ6L2 | 16 | -5.31 | 1.12E-07 | 1.02E-05 | -0.14 | 1.85E-03 | 0.07 | no |
| 13 | IGHV3-72 | 14 | -4.61 | 4.02E-06 | 2.99E-04 | -0.21 | 3.15E-03 | 0.30 | no |
| 14 | CD33 | 19 | 4.24 | 2.25E-05 | 1.15E-03 | 0.04 | 3.58E-04 | 0.71 | no |
| 15 | IGHG2 | 14 | -4.07 | 4.74E-05 | 1.99E-03 | -0.18 | 2.78E-03 | 0.30 | no |
| 16 | GALNT6 | 12 | -4.06 | 4.99E-05 | 1.99E-03 | -0.04 | 1.29E-05 | 0.67 | no |
| 17 | CLN5 | 13 | -4.05 | 5.10E-05 | 1.99E-03 | -0.05 | 1.27E-04 | 0.13 | no |
| 18 | NTRK1 | 1 | 3.76 | 1.73E-04 | 6.16E-03 | 0.12 | 1.69E-03 | 0.65 | no |
| 19 | ST8SIA3 | 18 | 3.61 | 3.07E-04 | 8.98E-03 | 0.05 | 4.95E-04 | 0.94 | no |
| 20 | CRIP1 | 14 | 3.60 | 3.18E-04 | 8.98E-03 | 0.05 | 2.64E-02 | 0.06 | no |
| 21 | LRP2 | 2 | 3.34 | 8.32E-04 | 2.00E-02 | 0.10 | 8.84E-03 | 0.97 | no |
| 22 | CD300A | 17 | 3.22 | 1.30E-03 | 3.04E-02 | 0.08 | 4.23E-03 | 0.23 | no |
| 23 | LGALS3 | 14 | -3.08 | 2.04E-03 | 4.40E-02 | -0.03 | 9.54E-04 | 0.19 | no |
| 24 | TXNDC16 | 14 | -3.05 | 2.27E-03 | 4.77E-02 | -0.09 | 6.37E-03 | 0.64 | no |

We compared these 24 AD candidate causal proteins to prior proteogenomic studies in brain, CSF, and plasma. Thirteen had been identified previously in brain^10^ (n=8) or CSF^19^ (n=10) studies, and one (PILRB) in plasma^20,21^ (Figure 4a; Figure S1; Table S14). The remaining 10 are novel, and of these, 5 were measured only by MS (GALNT6, IGHV2-70, IGHV3-72, PILRB, TXNDC16).

**Figure 4.**
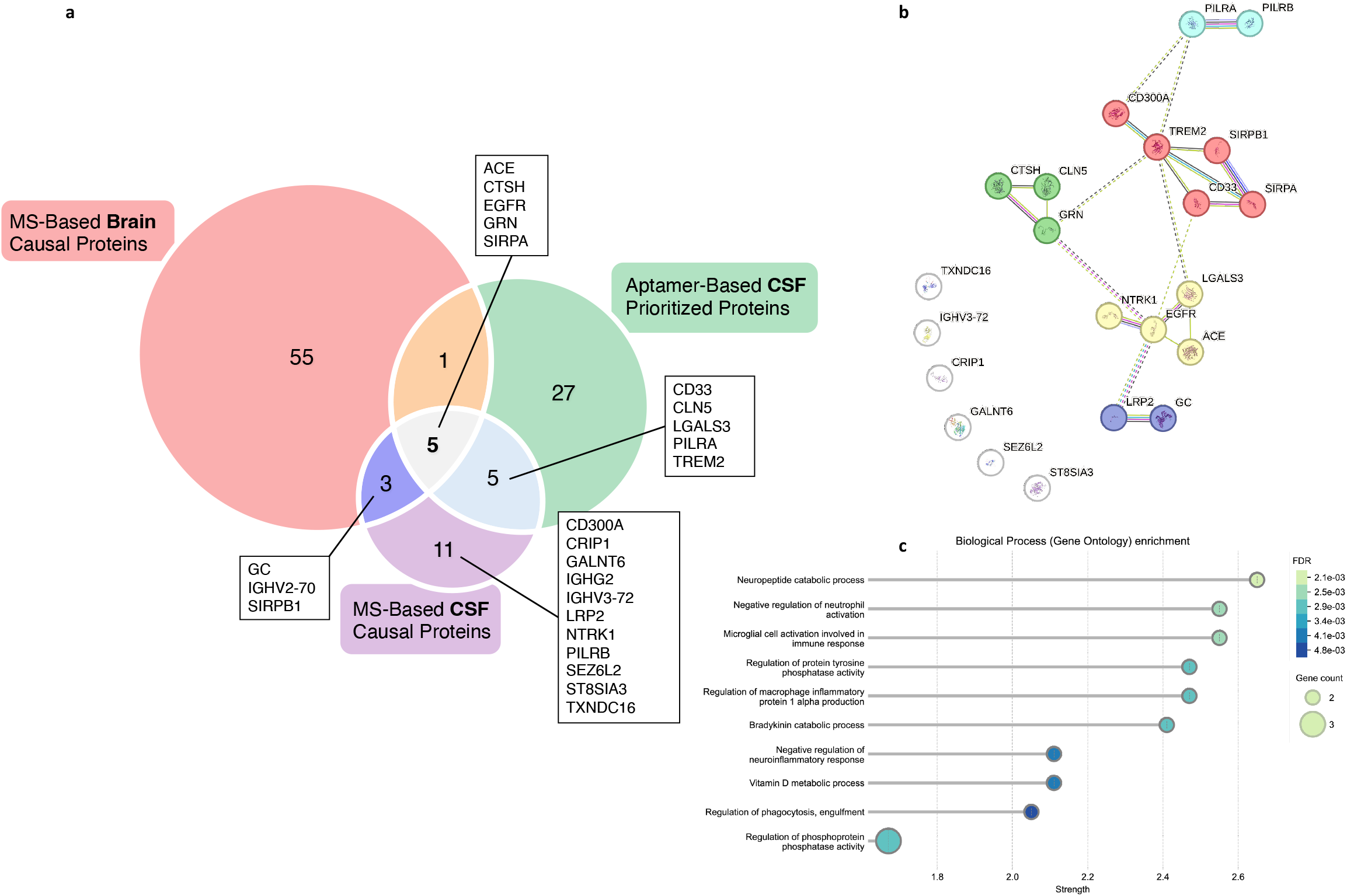
AD causal proteins and cross-tissue comparisons. **a**Comparison of AD causal proteins identified in brain and CSF using MS- and aptamer-based proteogenomics. The Venn diagram depicts proteins identified in published work (Wingo et al., 2025; Western et al., 2024) and in this MS-based CSF study. **b**. STRINGdb diagram for 24 AD causal proteins identified using MS-based CSF proteogenomics (two IGH proteins contained insufficient information and not depicted here – IGHV2-70 and IGHG2). Markov clustering reveals four functional modules comprised of a total 5 clusters known to contribute to distinct aspects of AD pathology. Clusters 1 and 2 (denoted by light blue and red) contain proteins that modulate microglia and macrophage activation^19-23^. Cluster 3 (yellow) contains proteins that regulate neurovascular growth and remodeling ^19, 24,26,27-28^. Cluster 4 (dark blue) are proteins governing vitamin D metabolism, which is implicated in many neuroinflammatory and neurodegenerative disorders. Cluster 5 (green) are proteins involved in proteolysis and lysosomal function^29-32^. The isolated nodes, include IGHV3-72 and the two additional immunoglobulin heavy chain components, suggesting a role for immunoglobulins in AD, while the remaining isolated nodes are best characterized in cell growth and carcinogenesis. **c**. Result of biological process enrichments highlight leukocyte activation, regulation of phagocytosis, and neuropeptide catabolic processes as the top 3 enriched FDR significant processes among the 24 causal AD proteins in CSF. The top three biological process enrichments by FDR highlight leukocyte activation, phagocytosis, and neuropeptide catabolism. In the context of our study, “leukocyte activation” and “regulation of phagocytosis” most strongly point to microglial activity, which is consistent with observed microglial dysfunction in AD^22,37-38^.

The 24 candidate AD causal proteins are involved in microglia and macrophage activation (PILRA, PILRB, CD300A, TREM2, SIRPB1, SIRPA, and CD33)^22–25^, regulating neurovascular growth and remodeling (LGALS3, NTRK1, EGFR, ACE)^26–30^, vitamin D metabolism (LRP2, GC), and proteolysis and lysosomal function (CTSH, CLN5, GRN, **Figure 4b-c**)^31–34^. Notably, three immunoglobulins (IGHV3-72, IGHG2, and IGHV2-70) implicate humoral immunity in AD risk (**Figure 4a**).

To assess whether candidate causal proteins show altered abundance in AD, we performed differential abundance analysis in CSF (ADNI, n=491) and brain (ROS/MAP, n=553). We found 297 proteins were differentially abundant in AD in CSF (FDR < 0.05; Table S15) and 3,038 proteins in brain (Table S16). In CSF, these proteins were enriched for synaptic, neurodevelopmental, and cell signaling processes (Tables S17). Of the 117 proteins differentially abundant in both tissues (Figure S2; Table S18), their genes were primarily enriched for synapse organization and neuronal function (Table S19).

Among the 24 CSF candidate causal proteins, none were differentially abundant in CSF, though two (CRIP1, TXNDC16) were in brain. Similarly, among 64 candidate causal proteins previously identified in brain^35^, none were differentially abundant in CSF, though 22 were in brain (Table S20).

## Discussion

Integrating deep MS-based CSF proteomics with the largest AD GWAS, we identified 24 candidate causal proteins for AD, 10 of which were not found in prior proteogenomic studies of brain, CSF, or plasma. The 24 AD candidate causal proteins converge on immune function, lysosomal biology, and neurovascular remodeling. Most notably, three immunoglobulin proteins (IGHG2, IGHV2-70, IGHV3-72) implicate humoral immunity, extending the role of the immune system from innate to humoral mechanisms in AD risk for the first time.

Eight candidate causal proteins were independently identified in brain^10^, suggesting they are the most robust findings. These proteins function along two major axes: neuroimmune activation (SIRPA, SIRPB1, CTSH, GRN, EGFR, GC) and proteostasis (ACE, GRN, and EGFR), with several proteins contributing to both. SIRPA and SIRPB1 regulate microglial activation and phagocytosis^36^, while GRN and CTSH contribute to lysosomal function and neuroinflammation^31–33^. GC regulates vitamin D bioavailability, which modulates neuroinflammation^37,38^. ACE showed the strongest negative association with AD risk; beyond its role in blood pressure regulation, ACE can convert Aβ42 into less pathological forms^28,39^. ACE, GRN, and EGFR can affect protein aggregation and clearance^28,33,39,40^.

The implication of humoral immunity in AD is novel. IGHG2 encodes the heavy chain constant region defining the IgG2 isotype, which preferentially recognizes polysaccharide antigens^41^. IGHV3-72 and IGHV2-70 are germline heavy chain variable segments whose allelic diversity can contribute to population level differences in humoral response^42^. IGHV2-70 was also identified as a candidate causal protein in brain, providing cross-tissue support. These findings suggest that variation in humoral immune function – not just microglial or innate immunity – contributes to AD risk

Beyond humoral immunity, 10 of 24 candidate causal proteins are involved in cell-surface immune recognition, including the three immunoglobulins as well as CD300A, CD33, PILRA, PILRB, SIRPA, SIRPB1, and TREM2. PILRA/PILRB and SIRPA/SIRPB1 represent two receptor families that fine-tune myeloid cell activation^22,23^. TREM2 is a well-known microglial receptor with a rare missense variant associated with AD^43^, and recent work suggests CD33 can inhibit TREM2-mediated phagocytosis in microglia^24,25^. Six proteins are linked to lysosomal function, including LGALS3, GRN, CTSH, CLN5, LRP2, and EGFR. The convergence of immune and lysosomal pathways is consistent with emerging models of AD pathogenesis in which impaired microglial clearance contributes to disease progression.

None of the 24 candidate causal proteins were differentially abundant in CSF, and only two (CRIP1, TXNDC16) were differentially abundant in brain. This apparent disconnect is expected. Mendelian randomization identifies proteins where genetic variation affects disease risk, potentially through subtle effects over a lifetime, whereas differential abundance reflects disease-state changes that may be downstream consequences rather than causes. This finding highlights that genetic causality and disease-state changes capture different aspects of protein involvement in AD.

MS-based proteomics quantified 1,066 CSF proteins not measured by affinity-based platforms, of which 5 are among the 10 novel candidate causal proteins. This shows the value of untargeted profiling for biological discovery. Cross-platform replication was moderate to high for cis pQTLs (*π*_1_ = 0.73 − 0.97) and lower for trans pQTLs (*π*_1_ = 0.42 − 0.85). Colocalization analysis found shared cis pQTLs between CSF and brain for 159 proteins, supporting CSF as a window into brain protein regulation for a subset of the proteome.

Interpretation of our findings should consider potential limitations of the study. Sample size may have limited our ability to detect lower abundant proteins and weaker pQTL signals or sex-biased effects on CSF pQTLs. This is mitigated by finding of moderate to high *π*_1_ rates between pQTLs for genes measured in other tissues and across platforms. Participants in our study are of European ancestry with high degrees of educational attainment. Our results ought to be tested for generalizability and future work would benefit from sampling across broader ranges of ancestries and ages. Additionally, the biological processes identified reflect those accessible to MS-based proteomics in CSF, and expanding tissue and technology coverage would likely provide a more complete picture of AD pathogenesis. We identified fewer APOE *trans* pQTLs than previously reported^13^, likely related to our covariate adjustment approach that included disease status. Whether APOE exerts a broad pleiotropic effect through direct changes in CSF proteins or indirectly through its influence on AD status remains an open question. Finally, the 24 AD causal proteins could be influenced by genetic pleiotropy, which could be addressed by experimental testing.

Our study has several strengths. TMT-MS is an untargeted, discovery-driven approach that does not require prior knowledge or the development of aptamers or antibodies for the proteins being profiled, enabling the identification of novel proteins. Our analytical strategy is rigorous, incorporating surrogate variable analysis to minimize potentially hidden confounding variables on pQTL mapping. Finally, we performed PWAS followed by Mendelian randomization with SMR/HEIDI to minimize the influence of LD, providing rigorous support for the 24 candidate AD causal proteins.

In summary, MS-based CSF proteogenomics identified 24 candidate AD causal proteins, including 8 that were previously identified in brain and 10 not found in prior studies. These proteins implicate lysosomal functions, neuroimmune regulation, and – for the first time – humoral immunity in AD pathogenesis, presenting promising new targets for future therapeutic development.

## Methods

### Study participants

#### ADNI

Data used in the preparation of this article were obtained from the Alzheimer’s Disease Neuroimaging Initiative (ADNI) database (adni.loni.usc.edu). The ADNI was launched in 2003 as a public-private partnership, led by Principal Investigator Michael W. Weiner, MD. The primary goal of ADNI has been to test whether serial magnetic resonance imaging (MRI), positron emission tomography (PET), other biological markers, and clinical and neuropsychological assessment can be combined to measure the progression of mild cognitive impairment (MCI) and early Alzheimer’s disease (AD). We used phenotypic, genetic, and proteomic data of participants recruited by ADNI. It enrolls participants aged 55 or older, who undergo a series of initial tests that are repeated at intervals over subsequent years, including a clinical evaluation, neuropsychological tests, genetic testing, lumbar puncture, and MRI and PET scans. ADNI has been approved by an Institutional Review Board.

#### ROS/MAP

Participants were recruited by the Religious Orders Study (ROS) or the Rush Memory and Aging Project (MAP), two ongoing longitudinal cohort studies of aging and dementia approved by an Institutional Review Board of Rush University Medical Center^44,45^. Both studies recruit older adults who enroll without known dementia and agree to annual clinical evaluation and brain donation at death. All participants signed an informed consent, the Anatomical Gift Act, and a repository consent to allow their study data to be repurposed. This study included 770 participants who had tandem mass tag (TMT) proteomics generated from the dorsal lateral prefrontal cortex (DLPFC), as described^46^, to compare with the ADNI MS-based CSF proteomics.

### Genetic data

ADNI genetic data was from array-based genotyping or whole genome sequencing data (WGS). For array-based genotyping, we performed quality control of the data using PLINK 1.9^47^ before performing imputation. We excluded subjects with an overall genotyping missingness > 10% or having mismatch between phenotypic and genotypic sex. For WGS data, we used Bystro^48^ to perform initial quality control, including determining 5 standard deviation outliers based on heterozygosity, silent replacement ratio, and genotyping completeness.

We restricted our genetic analysis to individuals with genetic similarity to European reference populations, as this was the only group with sufficient sample size for meaningful results in the ADNI dataset. To determine genetic similarity, we performed multidimensional scaling (MDS) analysis using PLINK 1.9, comparing individual-level genetic data to Phase 3 1000 Genomes European reference samples. For both array-based and WGS data, we included SNPs with MAF > 0.05 that passed LD filtering (100 Kb windows, 5 Kb steps, r^2^ < 0.15 for array data, r^2^ < 0.05 for WGS data). We excluded ambiguous variants (AT/TA/GC/CG) and retained only participants clustering within 6 standard deviations of the 1000 Genomes European reference populations.

We imputed array-based genotyping data using the TOPMed^49^ reference panel with hg38 coordinates following recommended quality control procedures: updating strand orientation, genomic positions, and reference/alternative allele assignments; removing A/T and G/C SNPs with MAF > 0.40; and excluding SNPs with differing alleles, allele frequency differences > 0.2, or absence from the reference panel. After imputation using TOPMed default parameters^49,50^, we retained SNPs with imputation quality R^2^ > 0.3. We then merged the imputed array data with WGS data from participants with genetic similarity to European reference populations, prioritizing WGS when both data types were available for the same individual. Post-merger quality control included removing variants with Hardy-Weinberg equilibrium deviation (p < 1×10^-7^), missing genotype rate > 10%, or MAF < 0.05. Finally, we excluded one individual from each pair of second-degree or closer relatives, retaining the individual with less genotyping missingness.

### Cerebrospinal Fluid Proteomics

#### Protein Digestion and Isobaric Tandem Mass Tag (TMT) Peptide Labeling

Baseline CSF samples from 1105 ADNI participants were digested as described before^51,52^. To normalize protein quantification across batches, a global internal standard (GIS)^53^ was created by aliquoting an equal volume (100 µL) of each CSF sample, post-digestion and HLB clean-up. All individual samples and GISs were dried using a speed vacuum concentrator (Labconco) and re-suspended in 100 mM TEAB buffer (50 μL). The TMTpro labeling reagents (5mg) were equilibrated to room temperature, and anhydrous ACN (200 μL) was added to each reagent channel (TMT reagent 16plex lot# YA357799; 134C and 135N lot# YB370079). Each channel was gently vortexed for 5 min, and then 10 μL from each TMT channel was transferred to the peptide solutions and allowed to incubate for 1 h at room temperature. The reaction was quenched with 5% (vol/vol) hydroxylamine (5 μL) (Pierce). All channels were then combined and dried by SpeedVac (LabConco). The combined sample was then resuspended in 500 μL of 0.1% TFA and then diluted 1:1 with 4% H3PO4 and desalted with a 30 mg MCX column (Waters). Samples were loaded onto the MCX column and then washed with 100 mM ammonium formate in 2% formic acid followed by a methanol and eluted with 300uL of 5% ammonium hydroxide in methanol. The eluates were then dried to completeness using a SpeedVac (LabConco).

#### High-pH Off-line Fractionation

Dried samples were re-suspended in high pH loading buffer (0.07% vol/vol NH4OH, 0.045% vol/vol FA, 2% vol/vol ACN) and loaded onto a Water’s BEH 1.7 um 2.1mm by 150mm. A Thermo Vanquish was used to carry out the fractionation. Solvent A consisted of 0.0175% (vol/vol) NH4OH, 0.01125% (vol/vol) FA, and 2% (vol/vol) ACN; solvent B consisted of 0.0175% (vol/vol) NH4OH, 0.01125% (vol/vol) FA, and 90% (vol/vol) ACN. The sample elution was performed over a 25 min gradient with a flow rate of 0.6 mL/min. A total of 192 individual equal volume fractions were collected across the gradient and subsequently pooled by concatenation into 96 fractions and dried using a SpeedVac.

#### Liquid Chromatography Mass Spectrometry

All fractions were resuspended in an equal volume of loading buffer (0.1% FA, 0.03% TFA, 1% ACN) and analyzed by liquid chromatography coupled to tandem MS. Peptide eluents were separated on Water’s CSH column (1.7um resin 150um by 15 cm) by a Vanquish Neo (ThermoFisher Scientific). Buffer A was water with 0.1% (vol/vol) formic acid, and buffer B was 99.9% (vol/vol) acetonitrile in water with 0.1% (vol/vol) formic acid. The gradient was from 3% to 35% solvent B over 17 mins followed by column wash and equilibration for a total of 23 mins. Peptides were monitored on an Orbitrap Astral spectrometer (ThermoFisher Scientific) fitted with a high-field asymmetric waveform ion mobility spectrometry (FAIMS Pro) ion mobility source (ThermoFisher Scientific). Two compensation voltages (CV) of −45 and −60 were chosen for the FAIMS. Each cycle consisted of one full scan acquisition (MS1) with an m/z range of 400-1500 at 120,000 resolution and standard settings and as many tandem (MS/MS) scans in 1.5 seconds. The Astral higher energy collision-induced dissociation (HCD) tandem scans were collected at 35% collision energy with an isolation of 0.5 m/z, an AGC setting of 100%, and a maximum injection time of 20ms. Dynamic exclusion was set to exclude previously sequenced peaks for 30 seconds within a 5-ppm tolerance window.

#### Database Search

The de-identified 6958 raw files (across 65 TMT-18 plexes) have been deposited at synapse.org with the SynID: syn59804727. These files were subsequently searched using FragPipe (version 22.0) as described^54–56^. The search was performed with a database of canonical Human proteins downloaded from Uniprot (20,402; accessed 02/11/2019), with added specific peptides for APOE E4 and E2 alleles, ABETA40, and ABETA42 peptides (total of 20405 sequences)^57^. The workflow used in FragPipe followed default TMT-16 plex parameters, used for both TMT-16 and TMT-18 experimental design (TMTpro). Briefly, precursor mass tolerance was −20 to 20 ppm, fragment mass tolerance of 20 ppm, mass calibration and parameter optimization were selected, and isotope error was set to −1/0/1/2/3. Enzyme specificity was set to semi-tryptic and up to two missing trypsin cleavages were allowed. Peptide length was allowed to range from 7 to 42 and peptide mass from either 200 to 5,000 Da. Variable modifications that were allowed in our search included: oxidation on methionine, N-terminal acetylation on protein, N-terminal acetylation on peptide along with off-target TMT tag modification on Serine, Threonine and Histidine, with a maximum of 3 variable modifications per peptide. Peptide Spectral Matches were validated using Percolator^58^. The false discovery rate (FDR) threshold was set to 1% and protein and peptide abundances were quantified using Philosopher^59^ for downstream analysis. The data from all batches were merged and protein levels were first scaled by dividing each protein intensity by intensity sum of all proteins in each sample followed by multiplying by the maximum protein intensity sum across all samples (i.e., column normalized). Instances where the intensity was ‘0’ were treated as ‘missing values’.

#### Quality Control

For CSF proteomic quality control, we kept proteins with data available in ≥ 200 subjects, selecting the highest abundance entry for proteins with multiple UniProt annotations. Protein abundances were normalized by sample-specific total protein content and log_2_-transformed. We checked for blood contamination by testing whether any sample was an outlier for hemoglobin subunits but found none. We excluded samples > 4 standard deviations from the mean of the first two principal components using iterative PCA. Linear regression was used to remove technical (batch) and biological (age, clinical diagnosis) effects from protein abundances. We then applied surrogate variable analysis (SVA package^60^) to identify and remove 67 additional confounding factors from the residual proteomic data. The final proteomic profiles were z-score standardized and limited to samples with available genetic data, resulting in 1006 subjects with 2949 proteins for pQTL mapping.

#### CSF pQTL mapping

We performed pQTL mapping using SNPs with MAF ≥ 0.05, defining cis pQTLs as variants within 1 Mb of the corresponding protein-coding gene (coordinates from BioMart^61^, accessed February 10, 2025) and trans pQTLs as variants outside this window. Linear models were fitted using PLINK^47^ with SNP genotype as the predictor and normalized protein expression as the outcome, adjusting for sex, 10 principal components, and genetic data source (WGS vs. array). Significance thresholds were p < 5×10^-8^ for cis pQTLs and p < 1.706 ×10^-11^ (5×10^-8^ / 2,931 proteins) for trans pQTLs. To address LD among significant pQTLs, we used PLINK’s ‘--clump’ function to identify independent index pQTLs with pairwise r^2^ < 0.1 within distance of 5 Mb.

#### CSF pQTL fine-mapping

We performed pQTL fine-mapping with SuSiE^15^ following its default parameters. SuSiE was designed to identify as many credible sets as the data support, each with as few variants as possible. For a given gene and its corresponding variants in the *cis* regulatory region, the output was the number of credible sets that had 95% probability of containing a variant with nonzero causal effect. We set the maximum number of credible sets for a gene to be 10 (default value).

#### CSF pQTL *π*_1_ rate

We compared our CSF pQTL findings (2,961 proteins, TMT-MS) with pQTLs from four studies: brain tissue (Wingo et al., n=1,013, TMT-MS)^10^, CSF (Western et al., n=3,506, SomaScan)^13^, and plasma (Ferkingstad et al., n=35,559, SomaScan; Sun et al., n=34,557, Olink)^5,6^. To assess replication, we estimated *π*_1_ (proportion of true associations) for each comparison by first calculating *π*_0_using the qvalue package, which takes the p-values from our index pQTLs when tested in each comparison study. Comparisons were made within each QTL type (i.e., cis to-cis or trans to-trans).

#### Integration of CSF proteomics and genetic data with AD GWAS results

We integrated CSF proteomic and genetic data with AD GWAS^18^ results using two complementary approaches. First, we performed PWAS using FUSION^4^, restricting genotypes to SNPs in the European LD reference panel. We estimated SNP-based heritability for each protein and declared those with p < 0.01 heritable. For heritable proteins, we trained prediction models (BLUP, LASSO, elastic net, BSLMM) using SNPs within 500 kb of the gene, retaining weights from the best-performing model. These weights were integrated with AD GWAS summary statistics to identify cis regulated proteins associated with AD (FDR < 0.05). Second, we performed Summary data-based Mendelian Randomization (SMR)^2^ on PWAS-significant proteins using pQTL and AD GWAS data to test whether CSF proteins mediate SNP-trait associations. We applied HEIDI^2^ testing to exclude associations likely due to LD (HEIDI p < 0.05). Proteins were considered causally implicated if they met all criteria: PWAS FDR < 0.05, SMR p < 0.05, HEIDI p > 0.05, and consistent effect directions on AD between PWAS and SMR analyses. All coordinates used GRCh38.

Comparing AD causal proteins identified in CSF between this and a published study^13^: Since the published study performed the PWAS for both *cis and trans* pQTLs (and not gene-based) using FUSION pipeline, and their cis and trans pQTLs were operationally defined as within 1MB window up and downstream of the gene, we compared our results with their results from cis pQTLs. Additionally, for genes with multiple somaIDs, we kept the top PWAS entry. In the 38 prioritized genes in the published study, 27 were results from cis pQTLs. We compared our CSF causal proteins with these 27.

#### STRING database

We use the STRING database^62^, which curates evidence from experimental data, computational predictions, text mining, and various databases, to examine protein-protein interactions (PPIs) encompassing physical PPI and functional associations. We input the 24 candidate AD causal proteins identified in CSF into STRING and examined their PPIs and biological pathways they are enriched in at FDR p<0.05.

#### Differentially expressed proteins in AD in both CSF and brain

We used available MS-based proteomic brain profiles from donors of the ROS/MAP studies as described^46^. To make the results comparable between brain and ADNI CSF proteomics data, we used clinically defined AD dementia as case and cognitively normal controls in both CSF and brain tissue. Regression models were fit for protein abundance adjusted for sex, age, and education. Proteins with FDR < 0.05 were considered significantly differentially expressed. AD diagnosis in ADNI was based on neuropsychological testing and a clinical exam with a neurologist at the time CSF was drawn for proteomic profiling.

Functional enrichment of differentially expressed proteins was performed using the clusterProfiler R package (v4.13.4)^63^. All proteins that passed quality control (n = 2,961) were used as the background set. Overrepresentation analysis was conducted using a one-sided Fisher’s exact test, and p-values were adjusted using the Benjamini-Hochberg false discovery rate (FDR) method. Gene Ontology (GO) terms with an adjusted p-value < 0.05 were considered statistically significant.

#### Examine expression of candidate AD causal proteins in CSF

Our prior work has identified 64 candidate causal brain protein in AD^10^. In the current study, we examined the overlap between these candidate causal proteins from brain and the proteins differentially expressed in AD in CSF.

## Data Availability

Raw mass spectrometry data and pre- and post-processed plasma protein abundance data and case traits related to this manuscript are available at https://www.synapse.org/Synapse:syn59804727 on the AMP-AD Knowledge Portal, which is a platform for accessing data, analyses and tools generated by the AMP-AD Target Discovery Program and other programs supported by the National Institute on Aging to enable open-science practices and accelerate translational learning. The data, analyses and tools are shared early in the research cycle without a publication embargo on secondary use. ADNI data are available on https://adni.loni.usc.edu/.

## Acknowledgments

We gratefully acknowledge the generosity of the participants and hard work of the ADNI study staff. We gratefully acknowledge the research volunteers and staff of ROS/MAP for their participation and contributions. We thank Jiaqi Liu for assistance in creating Figure 2.

## Resources

### Software

FragPipe (v22.0), MSFragger (v4.1), IonQuant (v1.10.27), diaTracer (v1.1.5), Philosopher (v5.1.1), Percolator (v3.6.5), R (v4.4.2), FUSION (commit e1ba5f7; https://github.com/gusevlab/fusion_twas), SMR (v1.02; https://yanglab.westlake.edu.cn/software/smr), susieR (v0.12.35; https://stephenslab.github.io/susieR/), SVA (v3.20.0; https://bioconductor.org/packages/sva/), and plink (v2.0; https://www.cog-genomics.org/plink/2.0/).

## Contributions

Conceptualization: APW, NTS, and TSW. Data Curation: AIL, APW, AS, DAB, DMD, EJF, FW, NTS, TSW, and ZM. Formal analysis: APW, ESG, MW, SMV, TSW, YL, and ZM. Initial Draft: APW, MW, TSW, and ZM. Supervision: APW and TSW. All authors revised and provided critical feedback on the manuscript and approved the manuscript.

---

**Supplementary Figure 1.**
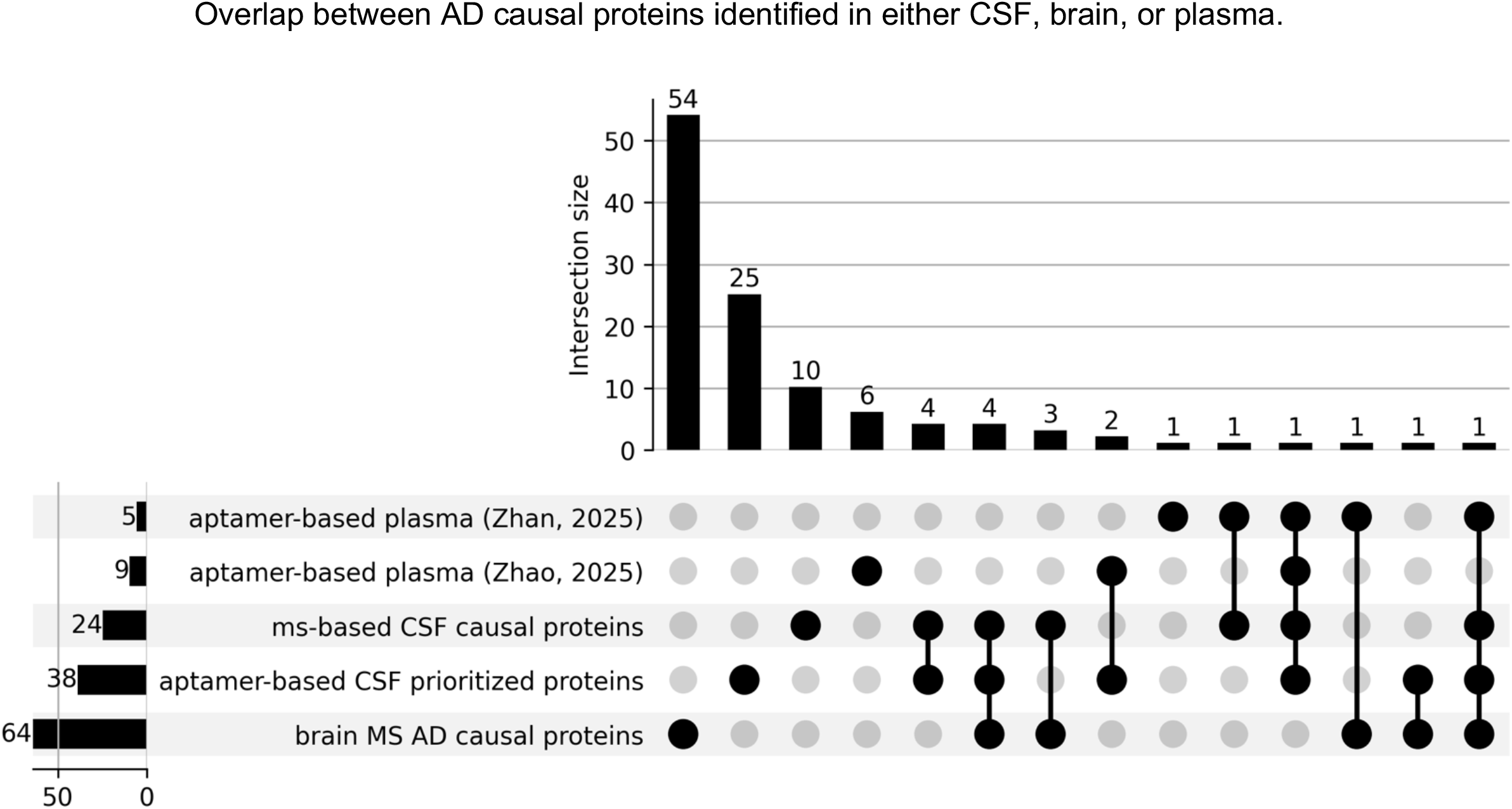
Comparison of AD causal proteins identified in CSF, brain, and plasma. Aptamer-based plasma study was from Zhan et al 2025 or Zhao et al 2025. MS-based CSF causal proteins are from the current study. Aptamer-based CSF prioritized proteins are from Western et al, 2024. Brain MS AD causal proteins are from Wingo et al, 2025.

**Supplementary Figure 2.**
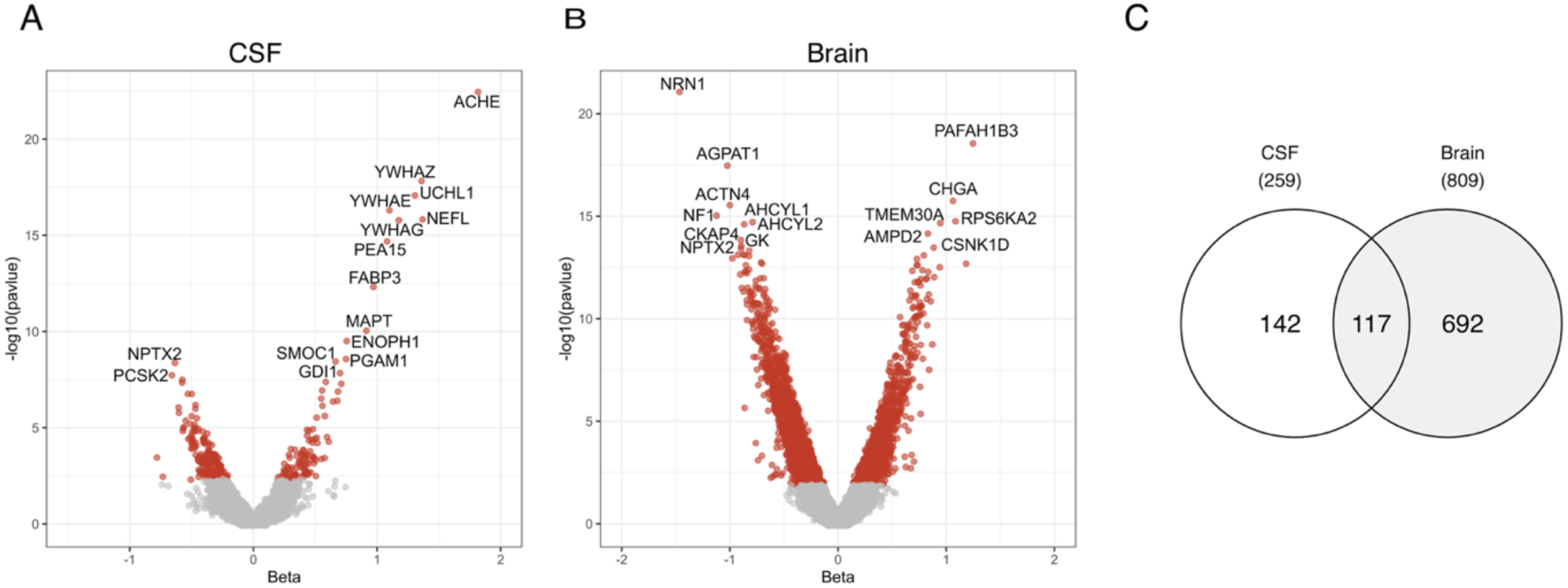
Comparison between CSF and brain proteins differentially expressed in AD dementia. **a**. Differential expression analysis of MS-based CSF proteome for AD dementia status in ADNI. **b**. Differential expression of MS-based brain proteome for AD dementia in ROS/MAP. **c**. Comparison between CSF and brain for proteins showing differential abundance in AD dementia. There were 2,961 proteins profiled in both CSF and brain, among those 117 were differentially expressed in AD dementia in both tissues. For the volcano plots in (a) and (b), red indicates differential expression at FDR <0.05 (significant) and black conveys not differentially expressed (FDR >0.05, non-significant). Beta-value was plotted on the x-axis and −log10 p-value on the y-axis.

